# Highly multiplex molecular inversion probe panel in Plasmodium falciparum targeting common SNPs approximates whole genome sequencing assessments for selection and relatedness

**DOI:** 10.1101/2025.03.07.25323597

**Authors:** Karamoko Niaré, Rebecca Crudale, Abebe A. Fola, Neeva Wernsman Young, Victor Asua, Melissa Conrad, Pierre Gashema, Anita Ghansah, Stan Hangi, Deus S. Ishengoma, Jean-Baptiste Mazarati, Ayalew Jejaw Zeleke, Philip J. Rosenthal, Abdoulaye A. Djimdé, Jonathan J. Juliano, Jeffrey A Bailey

**Affiliations:** Department of Pathology and Laboratory Medicine, Brown University, Providence, RI, USA; Center for Computational Molecular Biology, Brown University, Providence, RI, USA; Infectious Diseases Research Collaboration, Kampala, Uganda; Institute for Tropical Medicine, University of Tubingen, Tubingen, Germany; Department of Medicine, University of California, San Francisco, San Francisco, California, United States of America; Center for Genomic Biology, Institut d’Enseignement Supérieur de Ruhengeri, Ruhengeri, Rwanda; Noguchi Memorial Institute for Medical Research, University of Ghana, Legon, Ghana; Department of Pediatrics, HEAL Africa, Goma, Democratic Republic of the Congo; National Institute for Medical Research, Dar es Salaam, Tanzania; Department of Biochemistry, Kampala International University in Tanzania, Dar es Salaam, Tanzania; Department of Medical Parasitology, School of Biomedical and Laboratory Science, University of Gondar, Gondar, Ethiopia; Pathogens genomics Diversity Network Africa, Imm. Gwancoura, Sotuba, Bamako, Mali; Malaria Research and Training Center, University of Science, Techniques and Technologies of Bamako, Mali; Institute for Global Health and Infectious Diseases, University of North Carolina, Chapel Hill, NC, USA; Division of Infectious Diseases, School of Medicine, University of North Carolina, Chapel Hill, NC, 27599, USA; Department of Epidemiology, Gillings School of Global Public Health, University of North Carolina, Chapel Hill, NC, 27599, USA

## Abstract

**Introduction:** The use of next-generation sequencing technologies (NGS) to study parasite populations and their response and evolution to interventions is important to support malaria control and elimination efforts. While whole genome sequencing (WGS) is optimal in terms of assessing the entire genome, it is costly for numerous samples. Targeted approaches selectively enriching for sequence of interest are more affordable but sometimes lack adequate information content for key analyses.

**Methods:** We have developed a highly-multiplexed molecular inversion probe (MIP) panel (IBC2FULL) targeting single nucleotide polymorphisms (SNPs) with ≥ 5% minor allele frequency (MAF) in sub-Saharan African regions from publicly available *Plasmodium falciparum* WGS. We optimized the panel alone and in combination with antimalarial drug resistance MIPs in laboratory *P. falciparum* strains at different parasitemias, and validated it by sequencing field isolates from Democratic Republic of Congo, Ethiopia, Ghana, Mali, Rwanda, Tanzania and Uganda and evaluating population structure, identity-by-descent (IBD), signals of selection, and complexity of infection (COI)

**Results:** The new panel IBC2FULL consisted of 2,128 MIP microhaplotypes (containing 4,264 common SNPs) spaced by 5.1 - 18.4 kb across the entire genome. While these microhaplotypes were developed based on variation from sub-Saharan African WGS, 59.3% (2,529) of SNPs were also common in South-East Asia. The MIPs were balanced to produce more uniform and higher depth coverage at low parasitemia (100 parasites/μL) along with MIPs targeting antimalarial drug resistance genes. Comparing targeted regions extracted from public WGS, IBC2FULL provided higher resolution of local population structure in sub-Saharan Africa than current PCR-based targeted sequencing panels. Sequencing field samples, IBC2FULL approximated WGS measures of relatedness, population structure, and COI. Interestingly, genome-wide analysis of extended haplotype homozygosity detected the same major peaks of selection as WGS. We also chose a subset of 305 high performing probes to create a core panel (IBC2CORE) that produced high-quality data for basic population genomic analysis and accurate estimation of COI.

**Discussion:** IBC2FULL and IBC2CORE provide an improved platform for malaria genomic epidemiology and biology that can approximate WGS for many applications and is deployable for malaria molecular surveillance in resource-limited settings.

## INTRODUCTION

Malaria remains a significant public health problem with unacceptably high morbidity and mortality rates (World Health Organization, 2023). Despite substantial effort over the past decades, malaria control and elimination interventions are threatened by multiple factors. Currently, drug and diagnostic resistance (Ariey et al., 2014; Ashley et al., 2014; Dondorp et al., 2009; Uwimana et al., 2020; World Health Organization, 2023) threaten longstanding testing and treatment strategies in sub-Saharan Africa (SSA) where the vast majority of malaria deaths occur, primarily due to *Plasmodium falciparum (World Health Organization, 2023)*. Artemisinin partial resistance has emerged in multiple locations in Eastern Africa and is rapidly spreading (Conrad et al., 2023; Fola et al., 2023; Ishengoma et al., 2024; Juliano et al., 2023; Mihreteab et al., 2023; Uwimana et al., 2020; Young et al., 2024). Furthermore, effective vaccines inducing long-acting sterilizing immunity are lacking (RTSS Clinical Trials Partnership, 2014; RTS,S Clinical Trials Partnership, 2015), insecticide resistance continues to evolve and the mosquito species *Anopheles stephensi* has spread in SSA from Asia threatening urban environments, (Balkew et al., 2020; Ranson & Lissenden, 2016; Seyfarth et al., 2019). To support malaria elimination efforts by overcoming these issues, multipurpose genomic tools that can be deployed to cost-effectively achieve the molecular surveillance of the disease are currently needed.

The recent progress in next-generation sequencing technologies and analytical computational tools has paved the way for more effective malaria molecular surveillance to detect drug resistance markers, track malaria transmission, determine importation, and evaluate the impacts of interventions (Aydemir et al., 2018; Chang et al., 2017; Hathaway et al., 2018; Neafsey et al., 2021; Ngondi et al., 2017; Paschalidis et al., 2022; Rao et al., 2016; Schaffner et al., 2018; Volkman et al., 2012). Presently, single nucleotide polymorphisms (SNPs) across the genome are the most robust input for the molecular surveillance of malaria, readily allowing for population genetic measures and statistics as well as directly underlying nonsynonymous changes leading to drug resistance. Beyond direct assessment of drug resistance markers, current population genomic analyses of malaria parasite populations aim mainly but not only to i) understand origins, spatio-temporal spread and mechanisms of drug, diagnostic and vaccine resistances, ii) monitor parasite connectivity and importation into regions targeted for malaria elimination and iii) estimate complexity of infection (COI) to evaluate transmission intensity. To power these analyses, large numbers of variant sites from across the genome are optimal. Targeted sequencing techniques, which consist of sequencing the genome at specific loci, are generally inexpensive tools that can be used for large-scale molecular surveillance of malaria with numerous samples. Unfortunately, increasing the number of targets to be amplified together in a single tube for more robust analysis is currently a major challenge for methods such as multiplex PCR-based assays (Campino et al., 2011; LaVerriere et al., 2022; Tessema et al., 2022). As a result, existing affordable targeted sequencing tools cover a small proportion of variation across the genome and can not approach WGS level of genetic information useful for detecting selection and detailing identity-by-descent (IBD).

Molecular inversion probe (MIP) capture (Cantsilieris et al., 2017) coupled with next-generation deep sequencing was first applied in 2018 (Aydemir et al., 2018) and has now been used for malaria molecular surveillance particularly for large scale countrywide surveys of drug resistance and population structure (Conrad et al., 2023; Juliano et al., 2023; Moser et al., 2020; Verity et al., 2020; Young et al., 2024). This method is based on the utilization of short oligonucleotide probes to hybridize to and capture as single stranded circles regions of the genome. (Cantsilieris et al., 2017). The extension and ligation arms within a MIP (analogous to PCR primers) are homologous to sequences flanking the specific target. Unique molecular identifiers (UMIs) allow for error correction and accurate capture counts. After hybridization of the probes to the template, the targeted sequence between probe arms is captured by polymerase extension followed by end ligation. The closed circular DNA formed in this step is preserved during exonuclease digestion and thereby enriched. Later PCR amplification adds sample barcodes to create a sequenceable Illumina library. MIPs represent an ultra-multiplexed targeted sequencing technique that can be used to sequence thousands of loci in hundreds to thousands of samples on a single Illumina run. Beyond SNPs, MIPs also provide microhaplotypes that are more informative for relatedness and population structure analysis (da Silva et al., 2023; Tessema et al., 2022).

We sought to develop a new MIP panel that could better approximate the information content of whole genome sequencing (WGS). This new panel (IBC2FULL) captures microhaplotype information for 4,264 common SNPs from SSA densely distributed across the 14 chromosomes. We optimized the MIP panel and tested resulting SNPs to analyze IBD, population structure,COI, and selection metrics in field samples in SSA. We obtained an improved information content with estimates of population structure compared to existing PCR-based targeted sequencing tools as well as measures of IBD and selection signatures comparable to WGS.measures

## MATERIALS AND METHODS

### Identification of informative SNPs across the genome for probe design

To select SNPs for the design of our MIP panel, we harnessed whole genome sequencing read data from the Pf6 release (MalariaGEN et al., 2021) that was reanalyzed using an optimized GATK4 variant calling pipeline (Niaré et al., 2023). We selected SNPs detected from all field isolates collected in SSA that were bifurcated into West Africa, which was the majority of samples (N=2,164) and other African regions (N = 1,529). Other regions included East Africa (N=539), Malawi (N= 349) and Central West Africa (N = 275) and Central Africa (N=366). For the two groups, bcftools was used to retain SNPs with variant quality score log-odds > 0 and minor allele frequency (MAF) ≥ 5% within the core regions (excluding subtelomeric and hypervariable regions) (Miles et al., 2016) of the 14 chromosomes after removing loci and samples with > 10% and 20% missing genotypes, respectively and pruning based on linkage disequilibrium (LD) score ≥ 0.20.

### Panel design, optimization and data analysis pipeline

We used MIPTools (https://github.com/bailey-lab/MIPTools) to design probes to capture the targeted SNPs as previously described (Aydemir et al., 2018). Probes have two hybridization arms that are homologous to the flanks of target, and a nucleotide backbone linking them which includes priming sites for universal amplification to add sample indexes. The designed capture regions measure ∼100 nucleotides long. MalariaGEN Pf3k variant dataset (MalariaGEN et al., 2021) was provided to the algorithm to exclude probes that were sitting on SNPs and indels with MAF ≥ 5%. Paralogous regions with ≥ 98% sequence identity were excluded from the design as well as sequences with ≥ 98% identity with any region of the human genome to prevent non-specific capture.

The entire panel design, also called **<u>I</u>**nfectious Disease Epidemiology and Ecology Lab **<u>B</u>**ig SNP bar**<u>C</u>**ode 2 **<u>Full</u>** (termed IBC2FULL for short), was tested by pooling equal volumes of all MIPs (Fig 1A). After phosphorylation, this panel was used to capture laboratory strains (7G8, DD2 and HB3) in which human blood was spiked in before chelex extraction. Different probe pool concentrations (1μM, 2μM, 4μM and 8μM), parasitemias (100 parasites/μl, 1,000 parasites/μl, 4,000 parasites/μl and 10,000 parasites/μl) were used in this optimization. These were pooled after capture and barcoding and sequenced (Aydemir et al., 2018). MIPwrangler within the MIPTools suite built in the Jupyter Notebook were used for read processing and initial UMI-based PCR error correction and microhaplotype evaluation (Hathaway et al., 2018). Final processed microhaplotype data was formatted as FASTQ files and mapped onto the reference genome using BWA followed by variant calling into VCF using Freebayes (Garrison & Marth, 2012) from within MIPTools.

**Figure 1.**
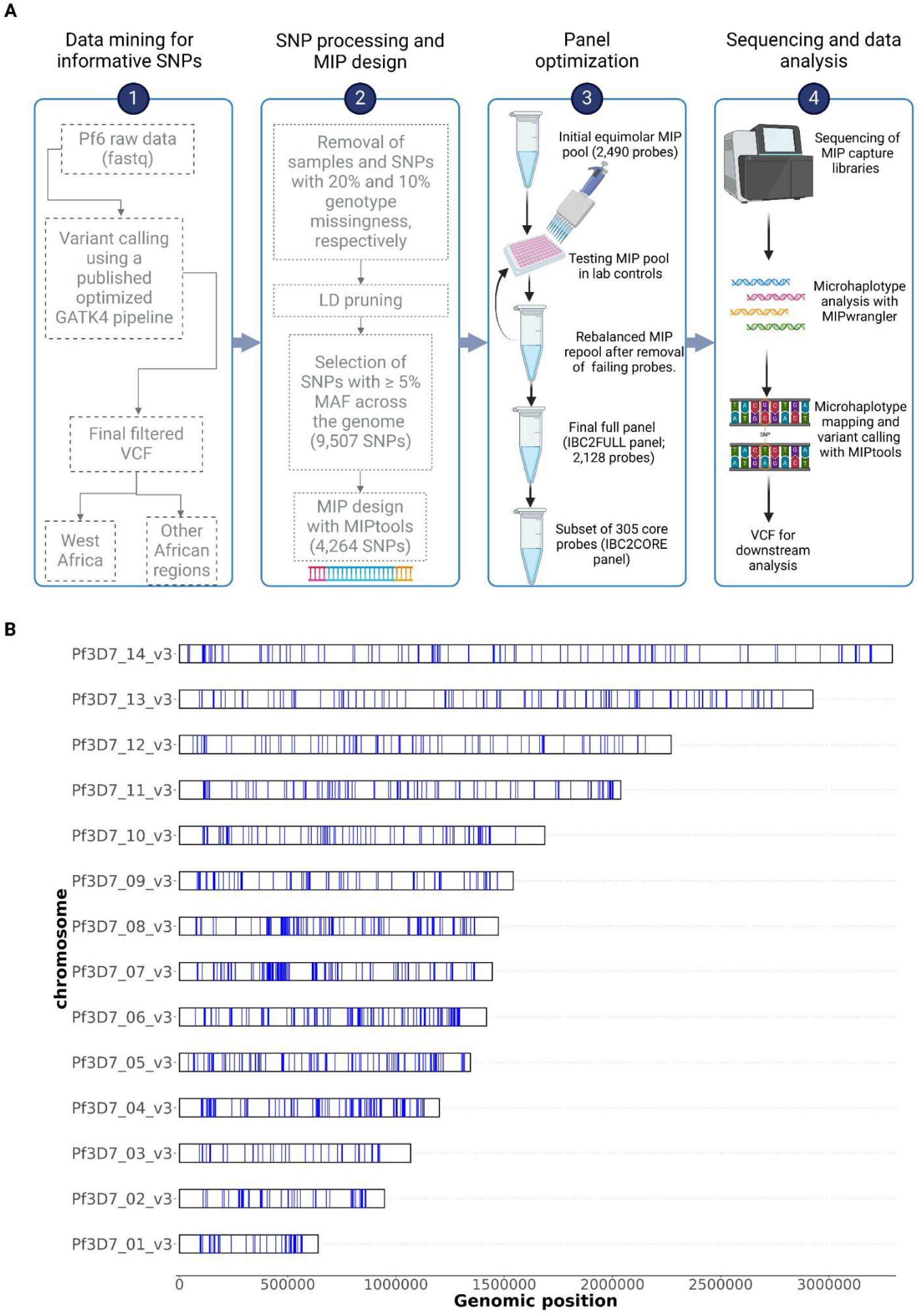
IBC2FULL panel design. **A)** Summary of IBC2FULL panel development pipeline. **B)** Genome-wide distribution of microhaplotype loci targeted by the IBC2FULL panel. Each blue band represents a microhaplotype captured by IBC2FULL. LD: linkage disequilibrium. MAF: minor allele frequency. VCF: variant call format. Pf6: Malariagen’s Pf6 release (MalariaGEN et al., 2021).

The sum rank of per probe UMI counts across samples at 1,000 parasites/uL parasitemia was used to estimate probe performance which allowed us to calculate the new volumes of probes to pool to create a final rebalanced panel. Probes that failed to provide at least 10X UMI depth per sample were not added in this final optimized panel.

For IBC2CORE inclusion, we selected a subset of 305 high-performing probes spaced on average at 100kb across the genome.

### Source of clinical samples used for MIP panel validation

We used archived dried blood spot and whole blood samples collected between 2020 and 2023 by seven collaborative studies in SSA, including Democratic Republic of Congo (DRC), Ethiopia, Ghana, MaliRwanda, Tanzania and Uganda. Ethical approval was obtained by each participating study with agreements for further use of the samples. Twenty high-quality samples lacking validated or candidate artemisinin partial resistance mutations were selected by country and subject to DNA extraction using Tween-Chelex or MagBead extraction.

### Population genomic analyses

#### Microhaplotype heterozygosity analysis

Genomic loci captured by the MIPs were considered as independent microhaplotypes and the total numbers of their unique sequences in our various dataset were estimated using custom scripts. All SNP positions with missing genotypes were removed before concatenating nucleotides from polymorphic sites to build haplotype sequences. Microhaplotype heterozygosity was computed using the formula 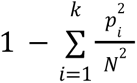 where p is the individual microhaplotype count and N the sum of all microhaplotype counts found at a given capture location.

#### Estimation of complexity of infection using MIP data

For VCFs generated from IBC2FULL, we only used targeted SNP positions for COI calculation. For WGS VCF, we used SNPs with MAF ≥5% that remained after variant recalibration and removal of sites and samples with ≥ 10% and 20% genotype missingness, respectively. *THE REAL McCOIL* package (Chang et al., 2017) was used to compute COI. The total numbers of Markov chain Monte Carlo and burn-in iterations were set to 2000 and 500, respectively.

#### Population structure analysis

To analyze the population structure, we pruned targeted SNP data based on minor allele frequency ≥ 1% (Amambua-Ngwa et al., 2023), LD < 0.20 and sample missingness rate < 20%. PLINK was used to compute the pairwise variance-standardized genetic relationship matrix. We used this matrix to perform and visualize principal component analysis using R packages PCA and factoextra, respectively. For validation, IBC2FULL was compared with MAD^4^HatTeR and SpotMalaria panels using their targeted loci extracted from Pf6 data.

#### Identity-by-descent analysis

Filtered MIP and WGS SNP data were used for pairwise IBD analysis using hmmIBD (Schaffner et al., 2018) with a maximum number of fit iterations of 5 assuming 0.1% genotype error rate. Sample pairs with at least 60 informative sites were selected for clustering and visualization using the igraph R package.

#### Analysis of selection signatures

The *rehh* R package was used to compute allele-specific integrated haplotype homozygosity score (iHS) across the entire core genome using filtered VCFs from IBC2FULL and WGS. To test whether an allele is under selection with significant increase in the iHS, the *P* value was calculated using the formula *P* = −log_10_(2Φ(−|iHS|)) where Φ (x) is the normal distribution function (Gautier & Vitalis, 2012).

## RESULTS

### IBC2FULL is an optimized panel approximating the information content of the whole genome

After a stepwise optimization (Supplementary Methods, Supplementary Table 1, Supplementary Figures 1, 2), we successfully developed a large barcode panel (IBC2FULL) of 2,128 MIPs for 4,264 SNPs representing 44.9% of common (≥ 5% MAF) SNPs found in SSA across the entire genome excluding *var* genes and hypervariable regions (Miles et al., 2016) (Figures 1A, B). However, 59.3% (2,529) of these informative SNPs were also found in South-East Asian parasite populations. The panel targeted diverse regions with a mean microhaplotype heterozygosity score of 0.4, 95% CI: 0.39 - 0.41 (Figure 2A).

**Figure 2.**
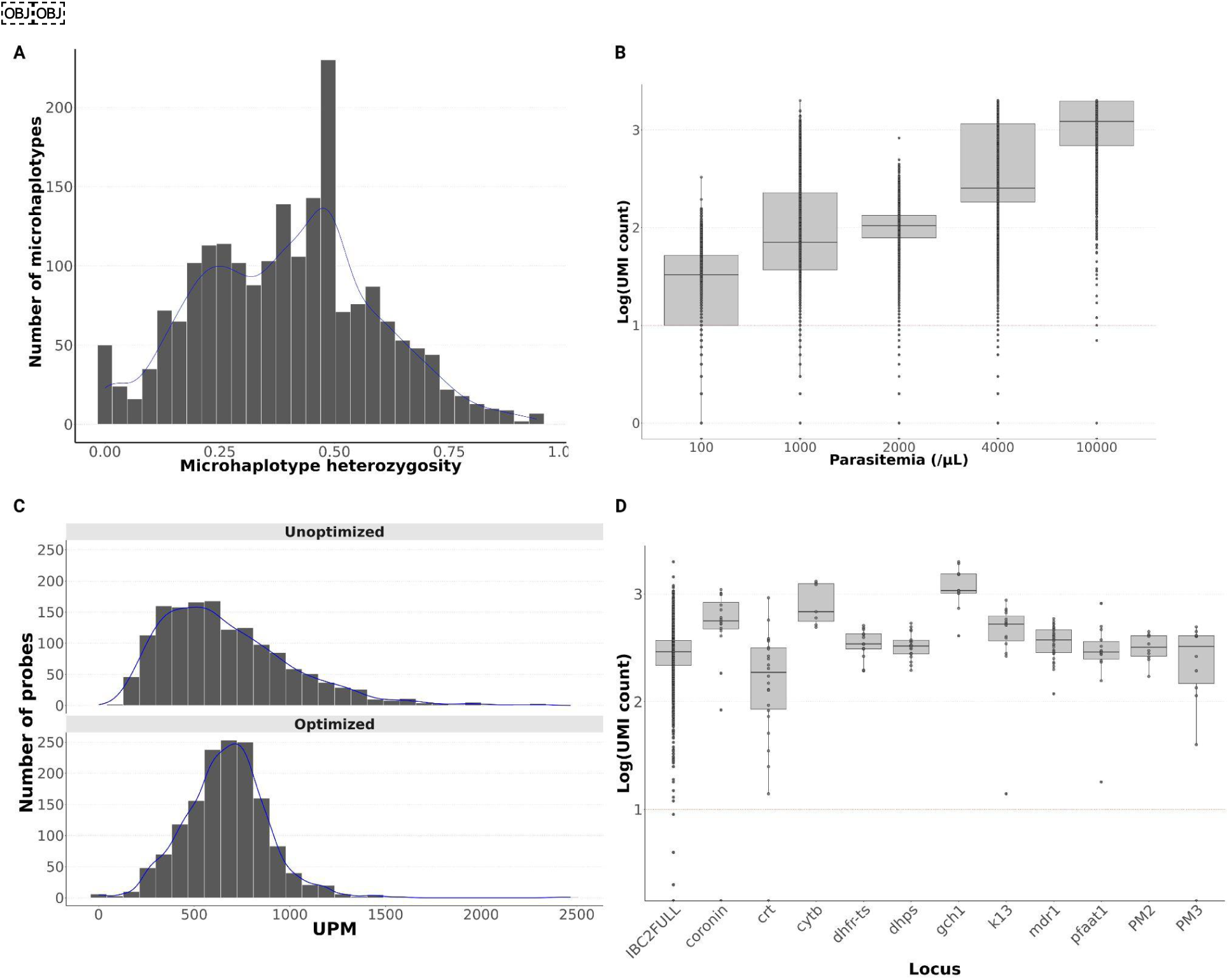
IBC2FULL panel optimization. **A)** Distribution of microhaplotype heterozygosity by probe. Heterozygosity was calculated from the Pf6 dataset in African field samples. **B)** Sequencing depth variation by parasitemia represented by log_10_ transformation of unique molecular index (UMI) count across samples. Each dot indicates the UMI count of one probe. **C)** UMI depth distribution of the unfiltered panel containing 2,490 probes after 1 and 2 Nextseq runs, and one run of the filtered and rebalanced panel with 2,128 probes remaining. Second Nextseq was done to verify if the low read depth of some probes is due to their intrinsic performance rather than insufficient sequencing. After this additional sequencing, 362 probes still showed low read depth and were removed from the final panel. UMI depth normalized per million per million (UPM). **D)** IBC2FULL and main antimalarial drug resistance genes showing similar sequencing depth (log10 transformation of UMI) after spiking drug resistance MIPs. The test was conducted in lab controls at 1,000 parasites/μL. 1,000 parasites/μL.

Read depth improved significantly with parasitemia (Figure 2B), but even lower parasitemia samples performed well with more than 10 UMIs per MIP on average at 100 parasites/μL. Despite this performance, there was an undesirable variability in the read depth across probes (standard deviation of per probe UMI count = 441) which was effectively addressed by re-pooling rebalanced volumes of individual MIPs (standard deviation = 77, Figure 2C). To ensure that IBC2FULL can be combined with other MIP panels without affecting the performance of both, we spiked in panel capturing antimalarial drug resistance genes that was previously used in field samples from Africa (Aydemir et al., 2018) and captured lab controls at 1,000 parasites/μL. After sequencing, no difference was found in read depth between IBC2FULL with and without the 11 genes tested with median UMI count > 100 (Figure 2D). To provide an even more targeted cost-effective panel (Supplementary Table 2), we selected 305 MIPs corresponding to 1 - 2 top performing probes based on UMI coverage in every 100kb window across the genome. This panel targeted 976 informative SNPs with high microhaplotype heterozygosity (0.435 on average), which represented 22.9% of IBC2FULL loci and 10.3% of the genome.

### SNPs captured by IBC2FULL display high-resolution population structure in sub-Saharan Africa

To evaluate the information content of IBC2FULL and IBC2CORE in terms of resolving local population structures, we performed principal component analysis with their targeted SNPs extracted from the Pf6 WGS from in sub-Saharan African samples and compared them to the full WGS core-genome SNP complement as well as two current targeted sequencing tools MAD^4^HatTeR (Aranda-Díaz et al., 2024) and SpotMalaria (Jacob et al., 2021). The first dimension explained up to 37.2% of variation between isolates and showed separation of West (Benin, Burkina Faso, Ghana, The Gambia, Guinea, Ivory Coast, Mali, Mauritania and Senegal) and Central West (Cameroon and Nigeria) African parasites from the rest of the populations with all tools except SpotMalaria. However, IBC2FULL displayed better discrimination resolution than other targeted tools and approximated the full SNP complement of WGS (Figures 3A, B, C, D, E). Together dimensions 1 and 2 further discriminated Central Africa (DRC) from East Africa (Ethiopia, Kenya, Madagascar, Tanzania and Uganda) and South-East Africa (Malawi) with IBC2FULL and WGS only. West African and Central West African parasites were also closely related but WGS and IBC2FULL showed the greatest population definition followed by MAD4HatTeR and IBC2CORE.

**Figure 3.**
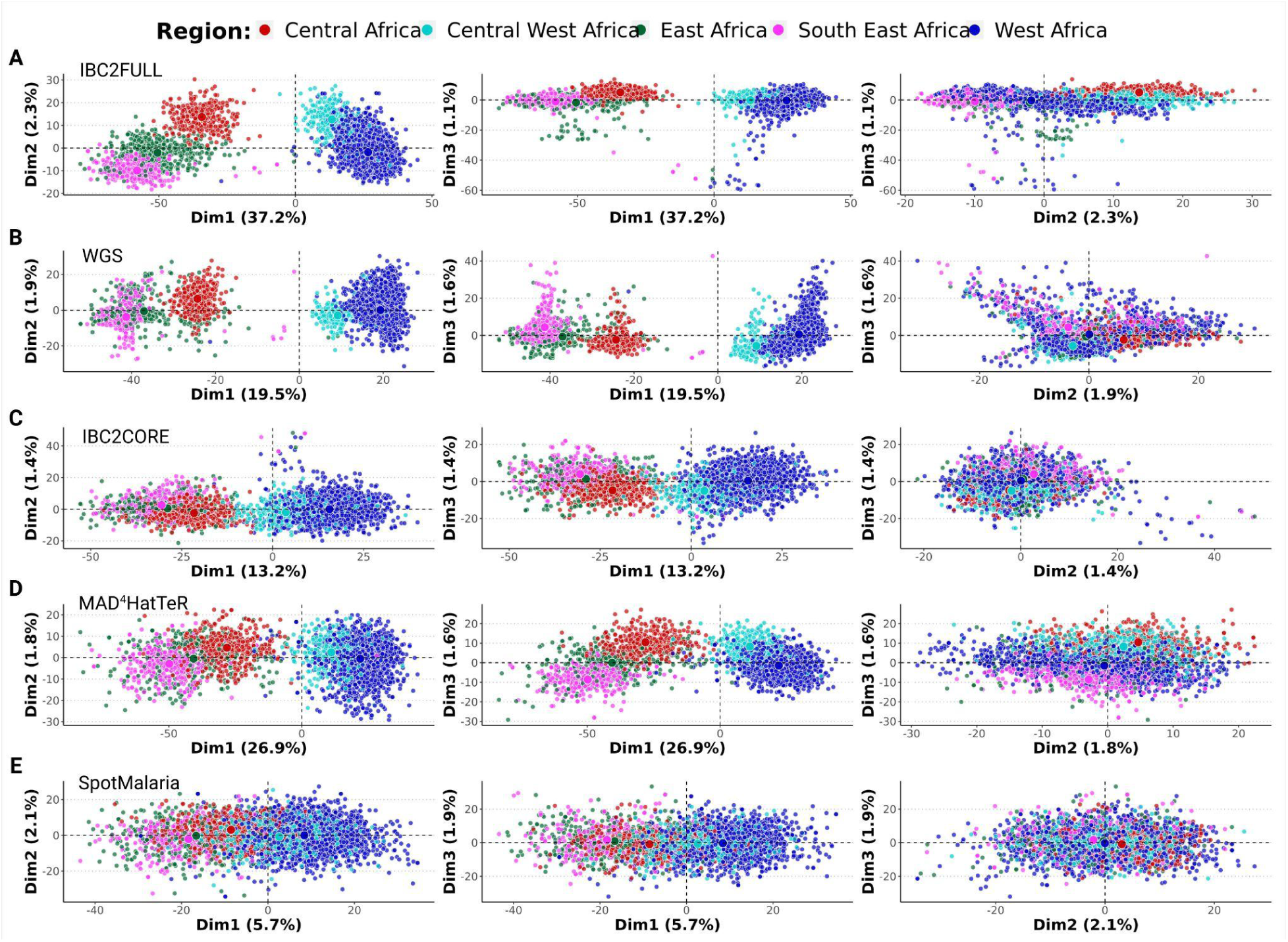
Local population structure in sub-Saharan Africa based on SNPs extracted from the Pf6 dataset. First 3 dimensions were displayed for **A)** IBC2FULL, **B)** Whole genome sequencing (WGS) after selecting SNPs with ≥ 1% minor allele frequency, **C)** IBC2CORE, **D)** MAD^4^HatTeR and **E)** SpotMalaria panels (n=3,693). The first three dimensions (Dim1, 2 and 3) of the PCA with the proportions of variance explained are shown. Each dot represents a sample colored by region defined in sub-Saharan Africa.

### IBC2FULL and whole genome sequencing equally detect relatedness and strong signatures of positive selection

To assess the ability of our IBC2FULL to accurately determine parasite relatedness, we used it to sequence field samples (n = 52) collected in Tanzania in 2022 for which WGS data was available (Ishengoma et al., 2024) and performed pairwise IBD analysis. We found a significant correlation (R = 0.81, P < 10^−16^) with the IBD sharing scores between IBC2FULL and WGS (Figure 4A) and no statistical difference in direct comparison of them (Figure 4B) although the latter is based on the comprehensive analysis of informative sites with ≥ 5% MAF. After classifying IBD sharing into short intervals of 0.1, there was a similar distribution of sample pairs across them with both tools with the majority of pairs below 0.1 IBD (Figure 4C). Additionally, there was a strong concordance of both methods in clustering field samples at IBD > 0.5 (Figure 4D).

**Figure 4.**
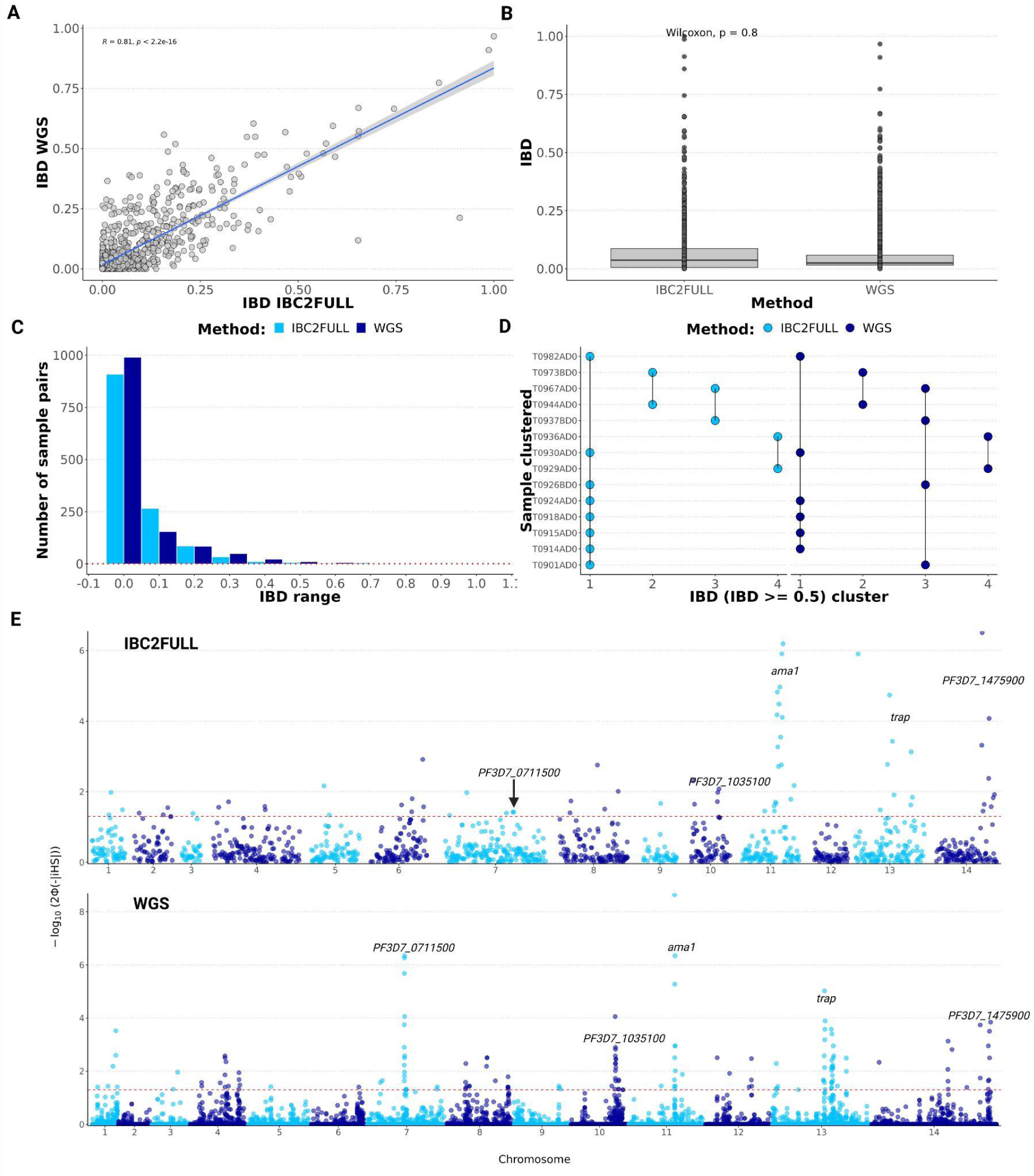
Measures of relatedness and signatures of positive selection across the genome in recent field isolates from Tanzania. **A)** Correlation of pairwise identity-by-descent (IBD) sharing between IBC2FULL and whole genome sequencing (WGS) (n = 52). **B)** Direct comparison of IBC2FULL and WGS pairwise IBD. **C)** Histogram showing sample pair distribution by IBD range for IBC2FULL versus WGS. **D)** Concordance between IBD-based sample clustering by IBC2FULL and WGS data. Four clusters were found in total by each of the tools at ibd ≥ 0.5. Each dot represents a sample assigned to a cluster name on the x-axis and the vertical lines link samples that belong to the same cluster. **E)** Manhattan plot showing *P* values (formula is shown on y-axis where Φ represents the Gaussian distribution function) of the integrated haplotype homozygosity score (iHS) across the 14 chromosomes of *Plasmodium falciparum* for IBC2FULL versus WGS (n = 52). Major signals detected by both tools were similar and are displayed including *ama1*, *trap,* PF3D7_0711500 (putative regulator of chromosome condensation), PF3D7_1035100 (unknown function) and PF3D7_1475900 (KELT protein). Each dot represents a SNP.

Because of the high SNP density of IBC2FULL, we were able to scan the entire genome for signatures of positive selection using the same data generated with field isolates from Tanzania. To test the null hypothesis of no selection, we estimated the *P* values around the iHS based on the Gaussian cumulative distribution for both WGS and IBC2FULL panels. The estimated SNP-level iHS score was equally normally distributed for both tools (Supplementary Figure 3), suggesting IBC2FULL can be used to detect strong signatures of positive selection accurately. Interestingly, IBC2FULL was able to detect all the strongest signals of selection (*P* <10^−4^) found with WGS, including *ama1, trap,* PF3D7_0711500 and PF3D7_1035100 (Figure 4E). Overall, we detected 24.3% of the WGS signals of selective sweep at *P* value <0.05, including mainly weaker selection (Supplementary Table 3).

### IBC2FULL and IBC2CORE are effective tools for measuring COI

We measured a strong correlation in the estimated COI between IBC2CORE and WGS and IBC2FULL (Figure 5A, *R = 0.94* versus WGS, *R = 0.97* versus IBC2FULL). IBC2FULL and WGS were also correlated (*R = 0.91*). For further validation, we conducted MIP sequencing on recent archived field samples (n = 140) from high transmission sites in West Africa (Ghana and Mali) and Central Africa (DRC) as well as lower transmission settings in East Africa (Ethiopia, Rwanda, Tanzania and Uganda) to assess COI. The prevalence of polygenomic infections ranged between 0% in Rwanda to 90% in Mali and was in keeping with the intensity of malaria transmission in these areas (Figure 5B). Having theoretically demonstrated that IBC2FULL loci clustered parasite populations by region defined in SSA with the same resolution as WGS using the Pf6 dataset (Figure 3), we confirmed these findings with our actual MIP data from recent samples (Figures 5C, D). Interestingly, both IBC2FULL and IBC2CORE displayed high-resolution population structure by country based on the first 3 principal components combined (Figures 5C, D).

**Figure 5.**
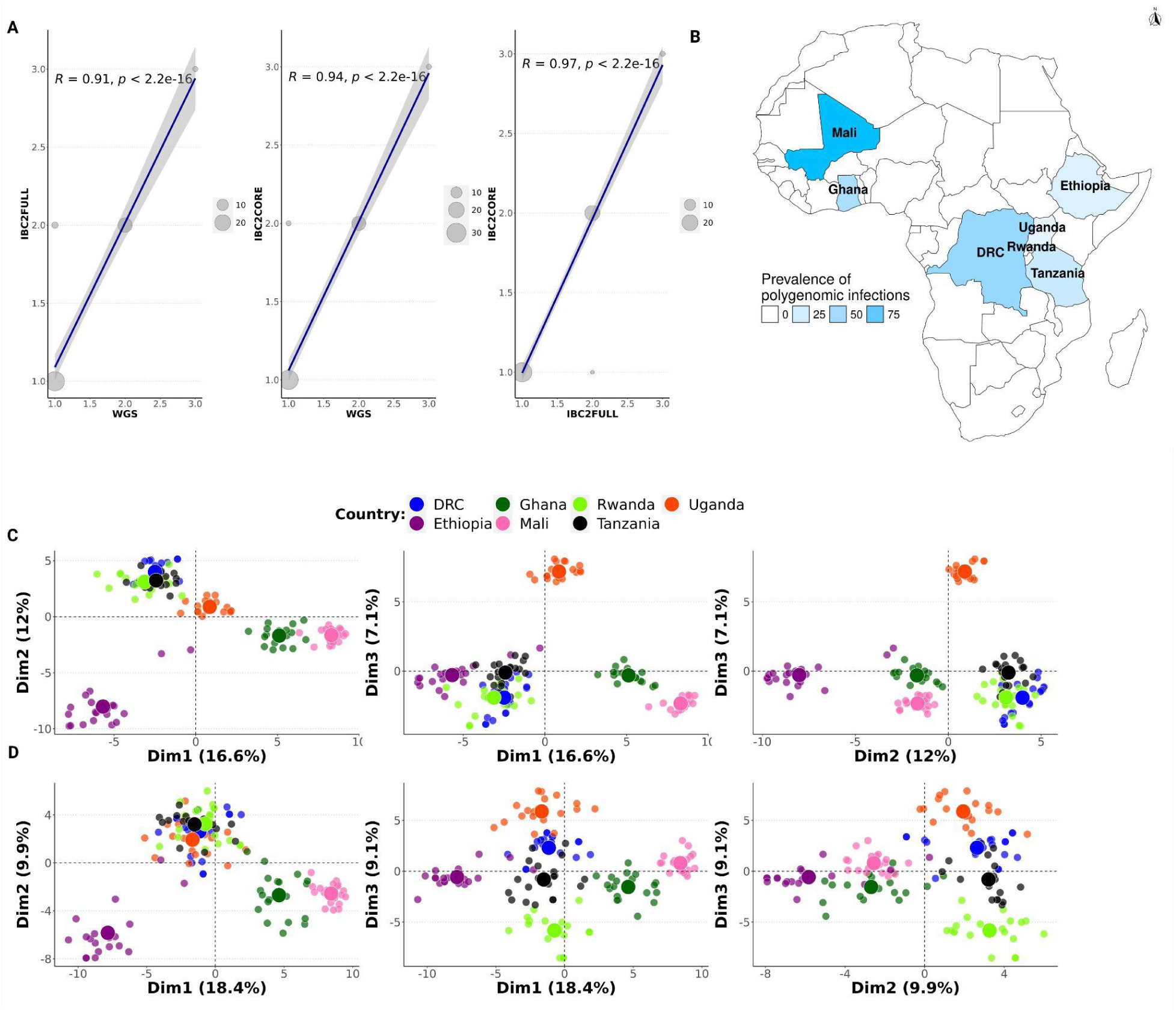
Complexity of infection and population structure in Africa based on molecular inversion probe (MIP) sequencing. **A)** Comparison of complexity of infection (COI) between IBC2FULL, IBC2CORE and WGS. Both x and y-axes show COI values but for different panels. Dot represent field isolates from Tanzania (n = 52) and dot size is proportional to the number of samples with the same COI. **B)** Prevalence of polygenomic infections (n = 140) in African countries studied using IBC2CORE. **C)** and **D)** Population structure in Africa based on MIP sequencing using recent field samples (n = 140) for IBC2FULL and IBC2CORE, respectively. First 3 dimensions are shown. Each dot represents a sample colored by country.

## DISCUSSION

While whole genome sequencing allows for comprehensive analysis of variants across the genome, only a subset of these are commonly used after filtering for quality and informativeness, especially those with high frequency in the circulating parasite populations. We successfully developed a new targeted sequencing panel IBC2FULL that incorporates ∼50% of the most common SNPs in African parasites. Comparison with WGS showed that we obtained the same levels of information for key population genomic metrics that are employed to inform malaria control and elimination efforts. Interestingly, we approximated the WGS in detecting strong signals of positive selection across the genome, which has not been previously evaluated for targeted panels.

The potential contributions of these new panels to malaria control and elimination and studying the biology of the parasites are numerous. IBC2FULL could refine measures of parasite connectivity within countries and across borders aiding in tracking the origins and evolution of drug or diagnosis resistances in regions where malaria parasite populations show minimal local population structure, such as much of SSA (MalariaGEN et al., 2021; Manske et al., 2012). Specifically, the high-resolution data generated with our panel could serve to resolve fundamental questions regarding the local emergence, importation, and clonal expansion of resistant parasites. It can be also used to monitor any parasite importation or disease outbreak in regions at the pre-elimination stage with very low prevalence of malaria. To assess the impact of control interventions, both IBC2FULL and its subset IBC2CORE, could be deployed to measure transmission intensity over time and across zones through the evaluation of the prevalence of polygenomic infections. With the possibility of scanning IBD and haplotype sharing across the genome, IBC2FULL can be leveraged to examine the extent and speed of the selective sweep due to drug resistance by analyzing SNPs that are located in the vicinity of molecular markers. Further selected WGS sequencing could be leveraged in combination with IBC2FULL to provide more powerful and defined resolution of selection.

Our findings showed that IBC2FULL provides higher resolution in population structure compared to existing amplicon sequencing tools, such as SpotMalaria and the microhaplotype panel MAD^4^HatTeR (Aranda-Díaz et al., 2024; Jacob et al., 2021). The improvement in resolution with IBC2FULL could be explained by the multitude of loci with increased microhaplotype heterozygosity covered, its SNP density across the genome, its representativity of the geography and good data quality with error correction. Our panels were validated using recent archived field samples from different regions in SSA and the relatedness and COI reflect the geographical locations and transmission intensities of the sampling sites, respectively (World Health Organization, 2023). With IBC2FULL, we reproduced the same results obtained from WGS in performing genome-wide scanning for signatures of positive selection and some of our hits with high iHS signals such as *ama1* and *trap* genes have been previously reported in Africa (Amambua-Ngwa et al., 2012; Coulibaly et al., 2022; Mu et al., 2010). Currently, the IBC2FULL panel requires only Illumina Nextseq platform and its total cost per sample was 20 times cheaper compared to WGS coupled with selective whole genome amplification using Illumina Novaseq X Plus (Supplementary Table 2). However, there is a minimum upfront investment of $19,675 in purchasing 10 μM of probes but this can be used to process 336,900 samples costing $3 each including capture, library preparation and sequencing. For obligatory upfront investment, the IBC2CORE panel costs only $2,820 and can be used to sequence 36,872 samples at $1.54 per sample.

Our full panel could be an effective alternative to WGS(Neafsey et al., 2008; Van Tyne et al., 2011), in terms of cost-effectiveness and throughput. Whereas WGS still remains mainly for fundamental research with a limited number of samples, the MIP technique has been already applied to monitor the spread of drug resistance in the context of molecular surveillance using thousands of field isolates (Juliano et al., 2023; Moser et al., 2020; Verity et al., 2020; Young et al., 2024). Our new panel is similar to these previous MIP sequencing panels in terms of laboratory protocols and computational data analysis and can be combined with them to analyze the prevalence of drug resistance markers and the genetic background of resistant parasites simultaneously to address multiple questions of interest. We demonstrated this by spiking independently designed probes for drug resistance (Aydemir et al., 2018) into IBC2FULL to extend the application of the panel without affecting the overall sequencing depth. As perspectives, the new development would be dedicated to increasing the resolution of EHH around drug resistance markers by densifying the panel with probes covering main drug resistance genes and flanking regions.

As a general limitation, MIP sequencing is currently considered as less sensitive compared to PCR amplicon sequencing methods (Aranda-Díaz et al., 2024; Aydemir et al., 2018; LaVerriere et al., 2022; Sadler et al., 2024; Tessema et al., 2022), which could hinder the use of our panels in low parasitemia samples below 100 parasites/μl. Currently, to mitigate this issue, one of the best approach is to perform selective whole genome amplification in low parasitemia samples prior to running IBC2FULL

In conclusion, we present a new cost-effective targeted sequencing tool that approximates WGS for many genomic analyses and fits well for malaria molecular surveillance using numerous field samples even in resource-limited settings.

## Data availability statement

The original contributions presented in the study are included in the article/Supplementary Material, further inquiries can be directed to the corresponding author.

## Ethics statement

This study did not include human and animal subjects. Participating studies received ethical approvals for future use of the samples.

## Author contributions

KN and JAB conceived the study. KN designed the panel, performed the data analysis and coordinated the laboratory assays. RC performed laboratory assays. JAB obtained funding and supervised the study. KN drafted the original manuscript. JAB and JJJ validated results and did major review and editing of the manuscript. AB, NWY, MC,PJR, VA,PR, AJZ, PG, JBM, SH, AG, AD, DI, JL, JP, JJJ contributed with data or samples shared through participating studies. All authors reviewed the article and approved the submitted version.

## Acknowledgments

We would like to thank all researchers and participants involved in the studies sharing data or samples.

