## Supplementary material for "Highly multiplex molecular inversion probe panel in Plasmodium falciparum targeting common SNPs approximates whole genome sequencing assessments for selection and relatedness": Supplemenary Materials

**Supplementary Methods:**

**IBC2 FULL panel development and optimization.**

Of 9,507 core genome-derived SNPs with ≥ 5% minor allele frequencies in sub-Saharan African parasite populations, we successfully developed a MIP panel for 4,264 (44.9%) requiring passing design quality criteria for both extension and ligation arms including lack of underlying variation, inadequate hairpin melting temperature, GC content and paralogy. We initially designed 2,490 probes but subsequently removed those that showed low performance in laboratory controls at 1,000 parasites/µL parasitemia. In total, 2,128 probes with a median insert size of 102.6 bp (Supplementary Table I) were retained in the full big barcode panel (IBC2FULL). Majority (76.9%) of variants targeted by the IBC2FULL panel had minor allele frequencies ≥ 10% (Supplementary Table I) with an average of 2 SNPs per probe, leading to high micro-haplotype heterozygosity scores. The targeted loci were densely distributed across each of the 14 chromosomes and spaced by only 5.1 - 18.4 kb (Supplementary Table I) with 45% of SNPs targeted by at least two different probes either on one strand or both.

For 100 parasites/µL parasitemia, the lowest we tested, a low panel performance was observed with reduced MIP pool concentrations (≤2 µM) overall where the UMI coverage fell below 10X at 1 µM (Fig. S1). There was a marked improvement in the UMI coverage per probe when the MIP pool concentration was increased to 4 µM and 8 µM. For 1,000 parasites/µL parasitemia, the panel performance was very high with UMI coverage > 10X for all MIP pool and DNA template concentrations. For 4,000 parasites/µL and 10,000 parasites/µL, high UMI coverages were also found but increasing the MIP pool concentration beyond 4µM showed no benefit. Overall, we found similar results at 0.01ng/µL, 0.025 ng/µL and 0.1ng/µL DNA template concentrations in which specific parasitemias were spiked into human blood before chelex extraction. At higher DNA template concentrations (5.36 ng/µL, 5.88 ng/µL and 6.88 ng/µL), we observed a reduced panel performance due to more human DNA outcompeting. To create the IBC2CORE panel, we ranked probes based on both the sum of UMI counts across samples at 1,000 parasites/µL parasitemia with 2 µM of MIP pool and microhaplotype heterozygosity. Top 1 - 2 performing probes were selected after every 100 kb to make a pool of 305 MIPs (IBC2CORE) showing higher sequencing depth compared to the remaining MIPs in the IBC2FULL panel (Fig. S2).

### Supplementary Tables

**Supplementary Table 1: Summary characteristics of the IBC2FULL panel.**

| Characteristics | MAF* ≥ 5% (i.e. all) | MAF = 5 - 10% | MAF ≥ 10% |
|---|---|---|---|
| Number of probes | 2,128 | 261 | 1,412 |
| Number of targets covered | 4,264 | 986 | 3,278 |
| Median insert size (bp) | 102.6 | 105 | 104.3 |
| 1st quartile insert size (bp) | 94 | 96 | 95 |
| 3rd quartile insert size (bp) | 113 | 114 | 113 |

*MAF = minor allele frequency.

**<u>Supplementary Table 2:</u> Cost comparison of IBC2FULL and IBC2CORE panels with whole genome sequencing**

| Panel | Probe order (10 µM each) | Number of MIP pools per order | Number of captures per pool | Number of captures per order | Cost per probe | Total cost per library (including probe cost for MIP) | Recommended Illumina platform | Sequencing cost per sample | Total cost per sample |
|---|---|---|---|---|---|---|---|---|---|
| IBC2FULL | $19,675.80 | 4 | 8,4225 | 336,900 | $0.06 | $1.26 | Nextseq | $1.83 | $3.09 |
| IBC2CORE | $2,820.07 | 4 | 9,218 | 36,872 | $0.08 | $1.28 | Nextseq/MiSeq | $0.26/$1 | $1.54/$2.28 |
| WGS | N/A | N/A | N/A | N/A | NA | $39.70 | Novaseq | $18.50 | $58.20 |

**<u>Supplementary table 3:</u>** Top WGS iHS signals compared to IBC2FULL.

| Gene name or ID | P value of iHS (IBC2FULL) | P value of iHS (WGS) |
|---|---|---|
| **AMA1** | **6.19261171000577** | **8.64151619952371** |
| **PF3D7_0711500** | **1.39305046250271** | **6.26192065291353** |
| **TRAP** | **4.73876106056855** | **5.02043117920354** |
| **PF3D7_1035100-PF3D7_1035200** | **2.0773087230096** | **4.05357505651282** |
| **PF3D7_1475900** | **6.50862353991685** | **3.84681930116264** |
| YIP1-HDA1 | 0 | 3.73830959381356 |
| SPECT1 | 0.875962690353979 | 3.57701998586976 |
| PF3D7_0114500 | 0 | 3.52108714682366 |
| ApiAP2-PMT | 0 | 3.40258195733424 |
| PF3D7_1448500 | 0 | 3.13233794526555 |

| | | |
|---|---|---|
| PF3D7_1475800 | 6.50862353991685 | 2.95469841611736 |
| PF3D7_1343800 | 0.468972876993136 | 2.95084999239442 |
| PF3D7_1450500 | 0.327982535364753 | 2.81415041696507 |
| PF3D7_1344100-HSP110 | 0.04082596147623 | 2.57636215299129 |
| PF3D7_0420600 | 0.390807225481692 | 2.57597684287277 |
| CelTOS | 0.81756239433197 | 2.50617024715144 |
| PF3D7_0820300 | 0.106145309528067 | 2.50617024715144 |
| PF3D7_1352900 | 3.12837086993673 | 2.4915717670038 |
| PF3D7_1238500 | 0 | 2.47289132447256 |
| PF3D7_1035200 | 2.0773087230096 | 2.45439246740621 |
| ApiAP2 | 0 | 2.41105491834461 |
| PF3D7_0421700 | 0.958012745746615 | 2.35220478172989 |
| PF3D7_1404800 | 0 | 2.33382678005934 |
| MPODD-PF3D7_0808500 | 0.565830901232849 | 2.28818634880647 |
| PF3D7_1301800 | 0 | 2.28818634880647 |
| K13-PF3D7_1343800 | 0 | 2.22072993534999 |
| PF3D7_0113300 | 1.98032866896627 | 2.18282383781193 |
| PGPS-PF3D7_0820300 | 0.106145309528067 | 2.18282383781193 |
| RAD5 | 0 | 2.18282383781193 |
| SPECT1-MyoA | 0.875962690353979 | 2.13501833473658 |
| PF3D7_0317300 | 0 | 1.96394441780174 |
| PF3D7_0425000-PF3D7_0425100 | 0 | 1.94407732372865 |
| CCHL | 0 | 1.92004165564063 |
| PF3D7_1137100-AEP | 0.129818174114432 | 1.88065096788422 |
| PF3D7_1343300-RAD5 | 0 | 1.84181889534949 |
| SURF8.2 | 0.576323245818273 | 1.79024139800907 |

| | | |
|---|---|---|
| PF3D7_1035200-GLURP | 0.740645293995203 | 1.67355055569287 |
| MSP7 | 0.68203581632375 | 1.66252266082035 |
| PF3D7_0705200 | 0.165644924998875 | 1.64877543538976 |
| PF3D7_0826100 | 1.11974511766134 | 1.63910690187572 |
| UT | 1.32969756894373 | 1.57594460380364 |
| PF3D7_0412200 | 0.557475171523276 | 1.57515239149321 |
| PF3D7_0419900 | 1.58142499063376 | 1.560593156319 |
| PF3D7_0425100 | 0 | 1.48034917366019 |
| EK-MGE1 | 0 | 1.441766441758 |
| PF3D7_1035000-PF3D7_1035100 | 2.0773087230096 | 1.441766441758 |
| PF3D7_1347600-ECT | 0.0341359267868758 | 1.441766441758 |
| pfa55-14 | 1.73742813018042 | 1.441766441758 |
| MSP1 | 0.216523716360091 | 1.43834301415561 |
| PF3D7_0412200-PF3D7_0412300 | 0.557475171523276 | 1.43834301415561 |
| PF3D7_0422000 | 0.84708215428982 | 1.43834301415561 |
| PF3D7_0809600 | 1.18020904125604 | 1.43834301415561 |
| PIP5K | 0 | 1.4365351115579 |
| PF3D7_1302700 | 5.90620252500942 | 1.42418632016112 |
| PF3D7_0511400 | 2.16363420911249 | 1.41460946168357 |
| PF3D7_0727500-PF3D7_0727600 | 0 | 1.41460946168357 |
| PF3D7_0100500 | 0 | 1.41008405651489 |
| CEPT-PF3D7_0628400 | 0.263068526824241 | 1.40330316099971 |
| PF3D7_1237100 | 0 | 1.4028876130329 |
| PF3D7_0104100 | 0.92669421634196 | 1.40252031007906 |
| SF3B2 | 0 | 1.39044193691006 |
| RPB12-PEPCK | 0.875962690353979 | 1.34995311552548 |

| | | |
|---|---|---|
| DNMT-PF3D7_0727400 | 0 | 1.32546498281408 |
| PF3D7_0713200-PF3D7_0713300 | 1.4245372331027 | 1.32420329199234 |
| PF3D7_0313600 | 0 | 1.32337939121129 |
| PF3D7_0930800-NFU1 | 0 | 1.32337939121129 |
| PF3D7_0713200 | 0.370125150775841 | 1.30142067009771 |
| RPS19 | 0 | 1.30142067009771 |

### Supplementary Figures

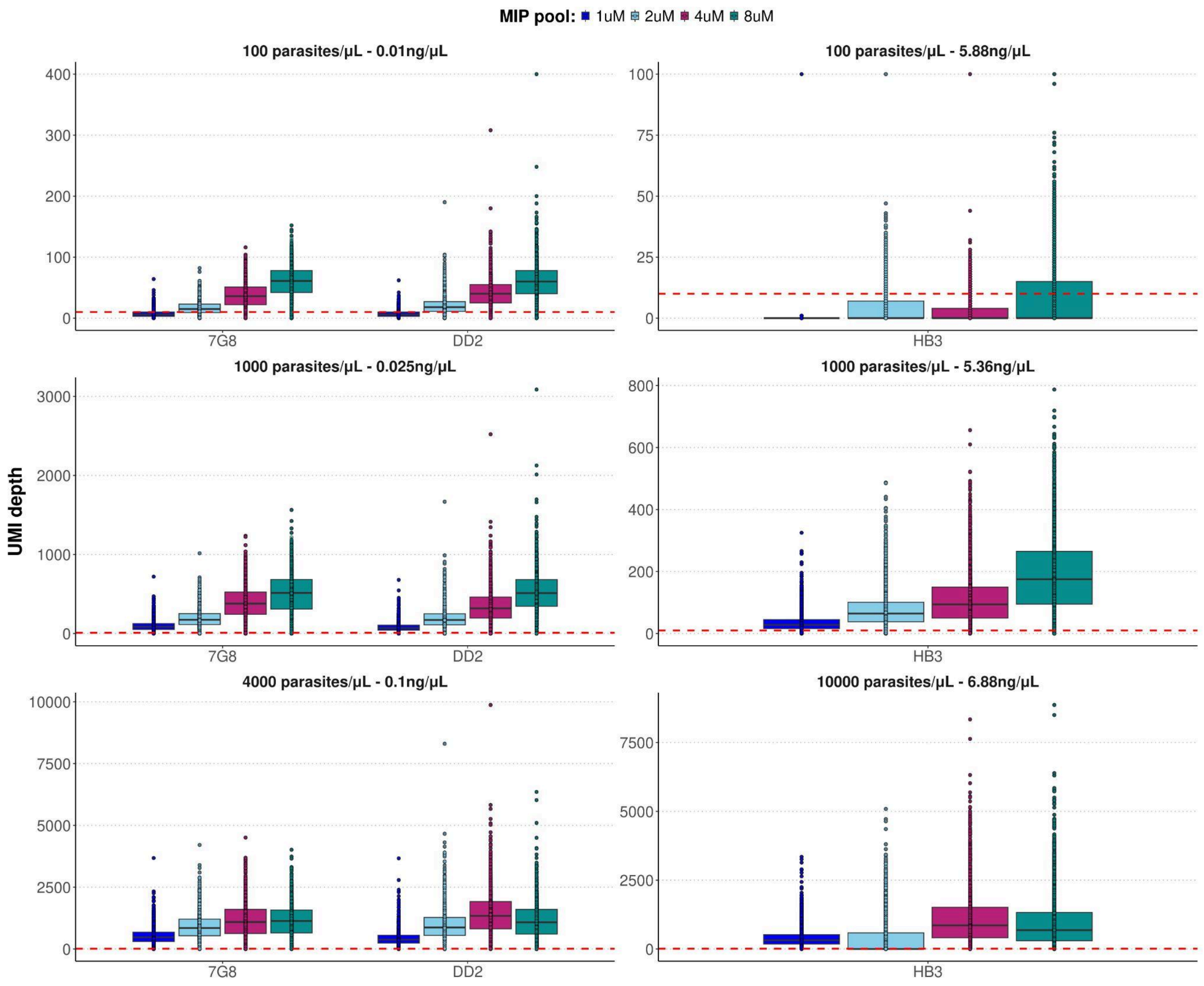


**Figure S1: Panel performance during the optimization.** The IBC2FULL panel was tested using the initial MIP tool concentration (8µM) and 2X (4µM), 4X (2µM) and 8X (1µM) dilutions at different parasitemias and DNA concentrations. The first row of the panel represents the optimization with the lowest parasitemia tested (100 parasites/µL)

at 0.01 ng/µL and 5.88 ng/µL of total DNA template. The second row corresponds to 1,000 parasites/µL parasitemia at 0.025 ng/µL and 5.36 ng/µL. The third row indicates high parasitemia optimization (4,000 parasites/µL at 0.1 ng/µl and 10,000 parasites/µL at 6.88 ng/µL). Three different lab strains (7G8, DD2 and HB3) were spiked into human blood at the desired parasitemias and used for the test After DNA extraction. The unique molecular index (UMI) depth was used to appreciate probe performance. The red dashed line represents 10X UMI which corresponds to reads coverage derived from 10 distinct captures of the targeted region.

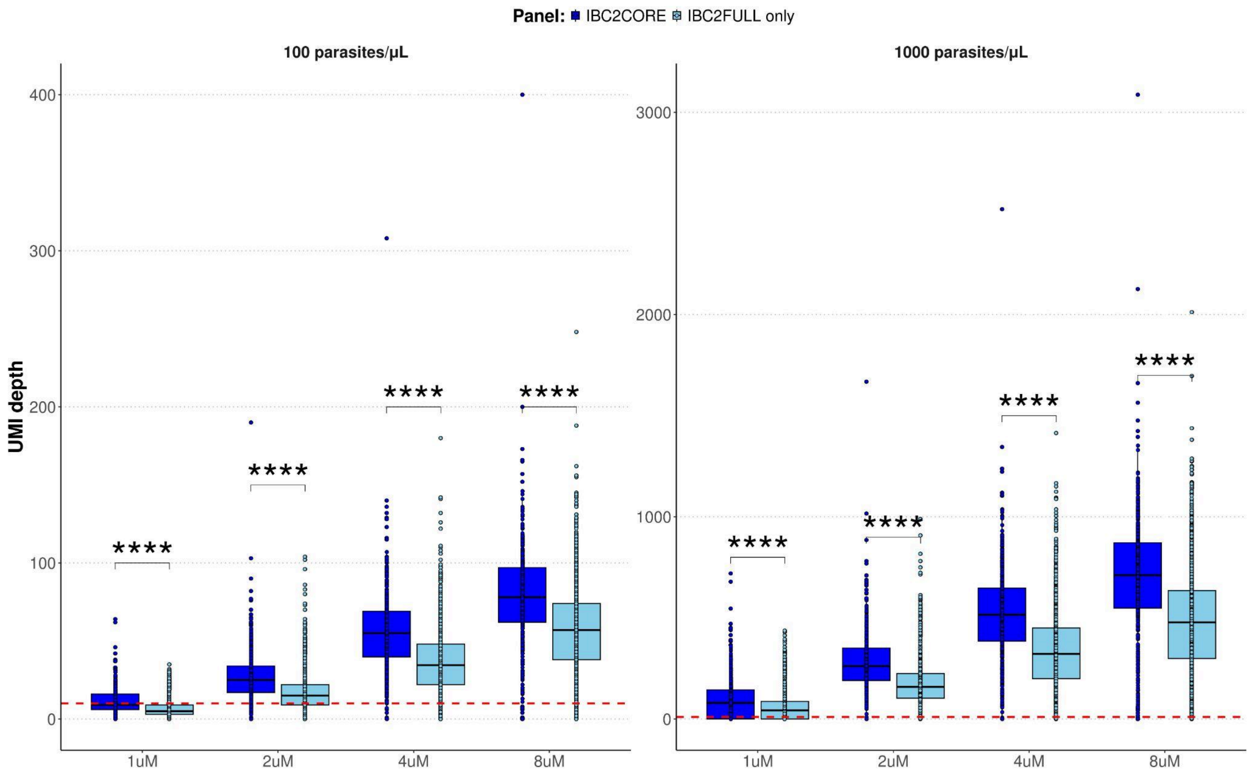


**Figure S2: Comparison of sequencing depth between IBC2CORE and remaining MIPs in IBC2FULL at 100 and 1,000 parasites/µL.** IBC2CORE was made of 305 MIPs and remaining MIPs in the IBC2FULL (IBC2FULL only) were 1823. Red dashed line represents 10 unique molecular index (UMI) counts. Stars indicate statistical significance ( $P<10^{-4}$).

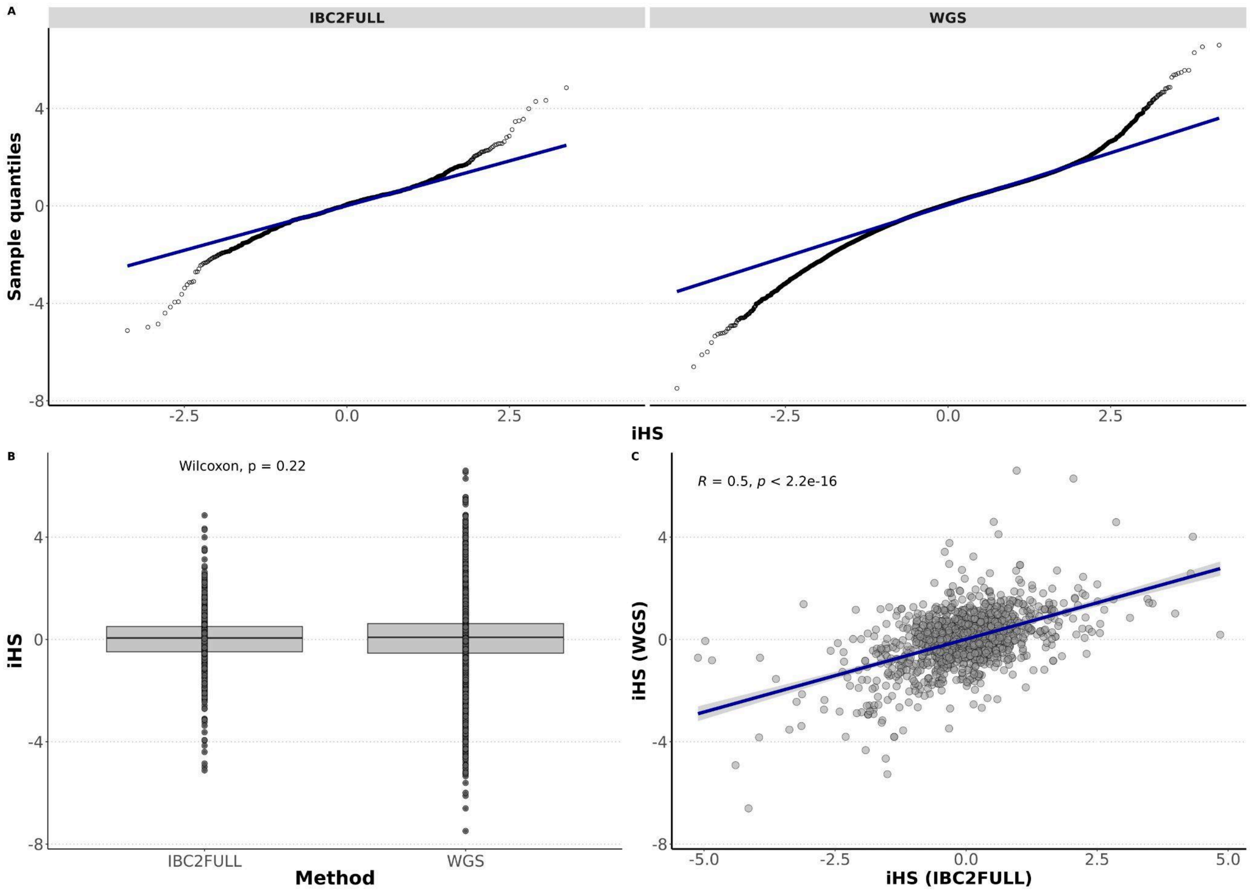


**Figure S3: Validation of the genome-wide analysis of extended haplotype homozygosity using IBC2FULL. A)** Q-Q plots of the integrated haplotype homozygosity score (iHS) for IBC2FULL versus whole genome sequencing (WGS) showing normal distributions. **B)** No significant difference between IBC2FULL and WGS in direct comparison of iHS. **C)** Correlation in iHS scores between IBC2FULLLL and WGS.

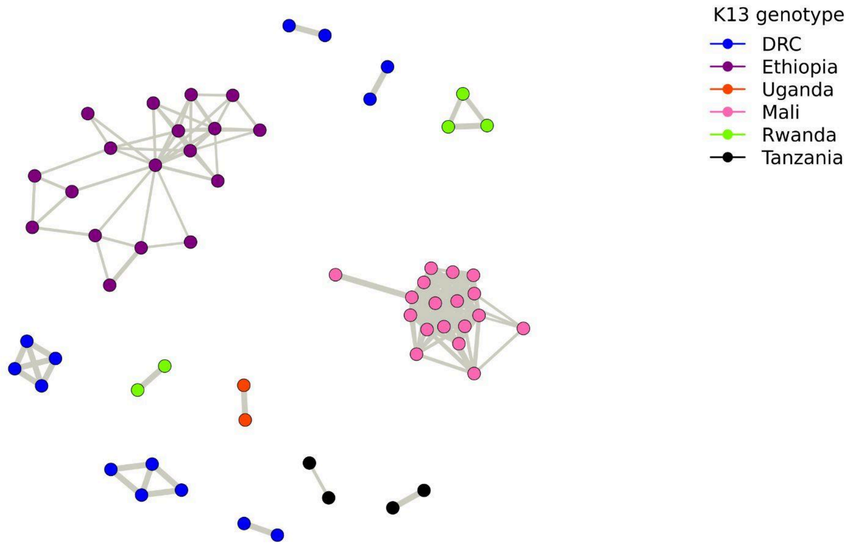


**Figure S4: Identity-by-descent (IBD)-based relatedness network of sub-Saharan African field samples (n=140) using IBC2FULL.** IBD ≥ 40 was used to draw the network.
